# Triggers for Freezing of Gait in Individuals with Parkinson’s Disease: A Systematic Review

**DOI:** 10.1101/2023.10.20.23297301

**Authors:** Carolina I. Conde, Charlotte Lang, Christian R. Baumann, Chris A. Easthope, William R. Taylor, Deepak K. Ravi

**Author notes:** Corresponding Author Dr. Deepak K. Ravi ETH Zürich Institute for Biomechanics Gloriastrasse 37/39 8092 Zurich Switzerland. **Financial Disclosure/Conflict of Interest:** Nothing to report.

## Abstract

**Background:** Freezing of Gait (FOG) is a motor symptom frequently observed in advanced Parkinson’s disease. However, due to its paroxysmal nature and diverse presentation, assessing FOG in a clinical setting can be challenging. Before FOG can be fully investigated, it is critical that a reliable experimental setting is established in which FOG can be evoked in a standardised manner, but the efficacy of various gait tasks and triggers for eliciting FOG remains unclear.

**Objectives:** This study aimed to conduct a systematic review of the existing literature and evaluate the available evidence for the relationship between specific motor tasks, triggers, and FOG episodes in individuals with Parkinson’s disease (PwPD).

**Methods:** We conducted a literature search on four online databases (PubMed, Web of Science, EMBASE, and Cochrane Library) using the keywords “Parkinson’s disease,” “Freezing of Gait,” and “triggers.” A total of 128 articles met the inclusion criteria and were included in our analysis.

**Results:** The review found that a wide range of gait tasks were employed in gait assessment studies on PD patients. However, three tasks (turning, dual tasking, and straight walking) were the most frequently used. Turning (28%) appears to be the most effective trigger for eliciting FOG in PwPD, followed by walking through a doorway (14%) and dual tasking (10%).

**Conclusions:** This review thereby supports the use of turning especially 360 degrees as a reliable trigger for FOG in PwPD. This could be beneficial to clinicians during clinical evaluations and researchers who wish to assess FOG in a laboratory environment.

## Introduction

Freezing of Gait (FOG) is a common disabling motor symptom that occurs in people with Parkinson’s disease (PwPD). In earlier stages of the disease, FOG is estimated to affect around 50% of PwPD, with the number increasing up to 80% as the disease progresses [1–3]. FOG is defined as “a brief, episodic absence or marked reduction of forward progression of the feet despite the intention to walk” [4]. In practice, it manifests itself in three different patterns: small ‘shuffling’ steps, alternating leg ‘trembling’ when stationary, and ‘akinesia’ with no discernible forward progression despite movement intention [4, 5]. The severity and complexity of these patterns can vary significantly from patient to patient and can even fluctuate within the same individual at different times [6]. FOG has a significant impact on mobility, frequently leads to falls, and reduces the quality of life for PwPD and their caregivers [7–10]. Additionally, FOG can lead to increased anxiety and insecurity while walking [10–12].

One of the prevailing hypotheses for the pathological mechanisms of FOG is the loss of automaticity, which is crucially reliant on both the basal ganglia and the brainstem [4, 13]. Freezing episodes that occur frequently in the OFF-medication state emphasize the significance of the dopaminergic pathway in relation to FOG [14–16]. However, FOG also occurs in the ON medication state, suggesting that a hypodopaminergic state only partially explains FOG [17]. Due to its inconsistency and its sensitivity to medication as well as emotional or environmental factors, further pathological mechanisms have been proposed [18]. Here, it has been suggested that FOG could be the result of a perceptual malfunction [19]. A study by Almeida and colleagues indicates that disturbed visuospatial feedback could explain why FOG often occurs when passing through doors or when approaching destinations [19]. Another study by Heremans et al. (2013) proposes that doorways may draw attention away from walking, or that approaching destinations may lead to the adaptation of locomotion, thereby eliciting FOG [20]. A further theory is that people experiencing FOG have insufficient inhibition to control unwanted movements, particularly when under time pressure or when required to shift between different response sets such as motor or cognitive tasks, indicating a possible frontal executive dysfunction, with or without enhanced levels of anxiety [20–23]. Overall, there is partial consensus that connectivity issues between the basal ganglia, the prefrontal cortex, and the frontoparietal areas could be the source of the problem causing FOG [14, 24, 25]. The presence of several different hypotheses for the causes of FOG demonstrates the complexity of its pathophysiology [5].

Assessing FOG in a clinical setting can be challenging due to the paroxysmal nature and diverse presentation of symptoms [26–28]. Multiple FOG-triggering settings have been associated with the condition, such as gait initiation [29, 30], turning [31–33], anxiety [34, 35], and walking in narrow spaces [36–38], among others. Three distinct phenomenological types of FOG have been identified in previous studies [5, 39]: asymmetric-motor, sensory-attention, and anxious freezing. Asymmetric motor freezing occurs mainly during turning, movement initiation, or when walking through narrow passages [5, 39–41]. Sensory attentive freezing is often a result of walking in the dark, walking through an unorganized space, or when the surface is sloped [5, 39]. Proprioceptive disturbances such as in concomitant polyneuropathy can contribute to freezing or mask it. Lastly, anxious freezing is triggered in stressful situations, such as when under time pressure or dual tasking [5, 26, 35, 42].

Most FOG episodes occur outside of a clinical environment as the awareness of being observed (Hawthorne effect) may enhance walking performance [43], making it difficult to study the symptom during a doctor’s visit or in clinical studies [42, 44–48]. Questionnaires such as the Movement Disorder Society - Unified Parkinson’s Disease Rating Scale (MDS-UPDRS) [49], the Freezing of Gait Questionnaire (FOG-Q) [50], or the New Freezing of Gait Questionnaire (NFOG-Q) [51] are available to confirm and determine the presence, frequency, and severity of FOG retrospectively [52]. However, due to recall bias accompanied by a cognitive decline patients experience difficulties when self-assessing FOG based on their perception [53]. Furthermore, minor differences in freezing severity may not be reliably detected by the NFOG-Q as it is a self-rated questionnaire and awareness of the freezing behaviour might change over time [53]. As a result, videotaped gait analysis is currently the standard for FOG detection in clinics [14]. During these analyses, patients often perform different gait tasks like turning or dual tasking, but there is a wide variation regarding the protocol and the tasks included in different studies and clinical examinations.

In order to better understand FOG and find effective therapies, many studies have attempted to recreate scenarios where FOG can be elicited in a reliable manner. Despite the availability of several questionnaires and analysis techniques to capture the appearance of freezing, the lack of coherent recommendations for reliably eliciting freezing episodes in both observational and interventional research has impeded progress in understanding FOG. This has led to sparse objective information on the detection and effectivity of treatments in clinical practice [5, 14, 24, 40, 54]. Some experimental studies have found certain walking tasks to be more effective in eliciting FOG, such as turning with a small radius or walking through doors [26, 55, 56]. Based on these findings, some research groups have developed obstacle courses that combine various triggers, such as turns and narrow passages, as well as straight walking or dual tasking elements [57, 58]. Additionally, virtual, or augmented reality technologies have been used to elicit FOG by providing individualized triggers and the ability to scale the difficulty and complexity of tasks [59–63]. Although there is a large collection of gait tasks used to assess FOG, the effectiveness of these individual triggers in eliciting FOG remains elusive [35, 64, 65]. Various studies, using various aforementioned triggers, were not able to elicit any freezing episodes during their protocol [19, 66, 67]. As a first step to guide further development of triggering paradigms, now the challenge is to ascertain which motor task serves as the most efficient trigger for FOG.

Therefore, to address this challenge, the aim of this study was to conduct a systematic review of the existing literature to evaluate the evidence linking specific motor tasks and triggers to FOG episodes in PwPD. By gaining a comprehensive understanding of these triggers, we aim to lay the foundations for advancing current knowledge of the pathophysiological mechanisms underlying FOG, optimizing treatments, and enabling the development of new therapies.

## Methods

### Search strategy and selection criteria

In October 2021, a literature search was carried out following the guidelines of the preferred reporting items for systematic reviews and meta-analysis (PRISMA) [68]. Four online databases, namely PubMed, Web of Science, EMBASE, and Cochrane Library, were searched to include only studies that reported FOG in PwPD during a walking task. The search was conducted on articles published between 1998 and 2021 and was restricted to original articles published in English in a peer-reviewed journal. The complete search string is available as a supplementary material (Supplementary Methods 1). Furthermore, the complete review protocol has been registered in the PROSPERO database (CRD42022330511).

### Selection of studies and data extraction

The process of selecting studies was aided by the EPPI-Reviewer 4 (V.4.12.5.0, EPPI-Centre, UCL Institute of Education, University of London, London, UK) and EndNote (EndNote 20, Clarivate, Philadelphia, US) software programs. Once duplicates were eliminated, two reviewers (CC, DKR) independently screened the titles, abstracts, and full texts, resolving any discrepancies through consensus. Two studies reported only the median value of FOG outcomes (average FOG count, FOG duration and percentage time with FOG), in both studies the median value was used as the mean.

A spreadsheet (MS Excel, version 2018, Microsoft Corporation, Washington, US) was used by one reviewer (CC) to extract the following information:

- Publication details such as author names, title, and publication year
- Study information including the number of individuals in freezing, non-freezing, and healthy control groups, gait tasks performed, and the technology / assessment scores used for FOG assessment
- Participant demographics such as age, gender, disease duration, disease stage, medication, and clinical assessments (MDS-UPDRS score [52] and Hoehn and Yahr score [61])
- FOG outcomes such as total FOG count, average FOG count, percentage of time spent frozen, etc.

If any information was missing or needed clarification, the authors of the included studies were contacted for additional details.

### Data Synthesis

In order to assess the most effective triggers, the data of all studies that stated the gait task triggering FOG were extracted and normalized by the number of studies reporting the same trigger. For every activity, the numbers of FOG episodes were summed and normalized by the number of studies reporting FOG episodes triggered by the respective task. Plots were created in the RStudio Software (R version 4.1.3, R Core Team, Vienna, Austria) using the ggplot2 package (version 3.4.2) [69].

## Results

### Study selection and Characteristics

After an initial search that yielded almost 4000 articles, 1600 were identified as duplicates. Following a screening of titles and abstracts, 486 articles remained eligible, and 128 of them met the inclusion criteria (Figure 1).

**Figure 1.**
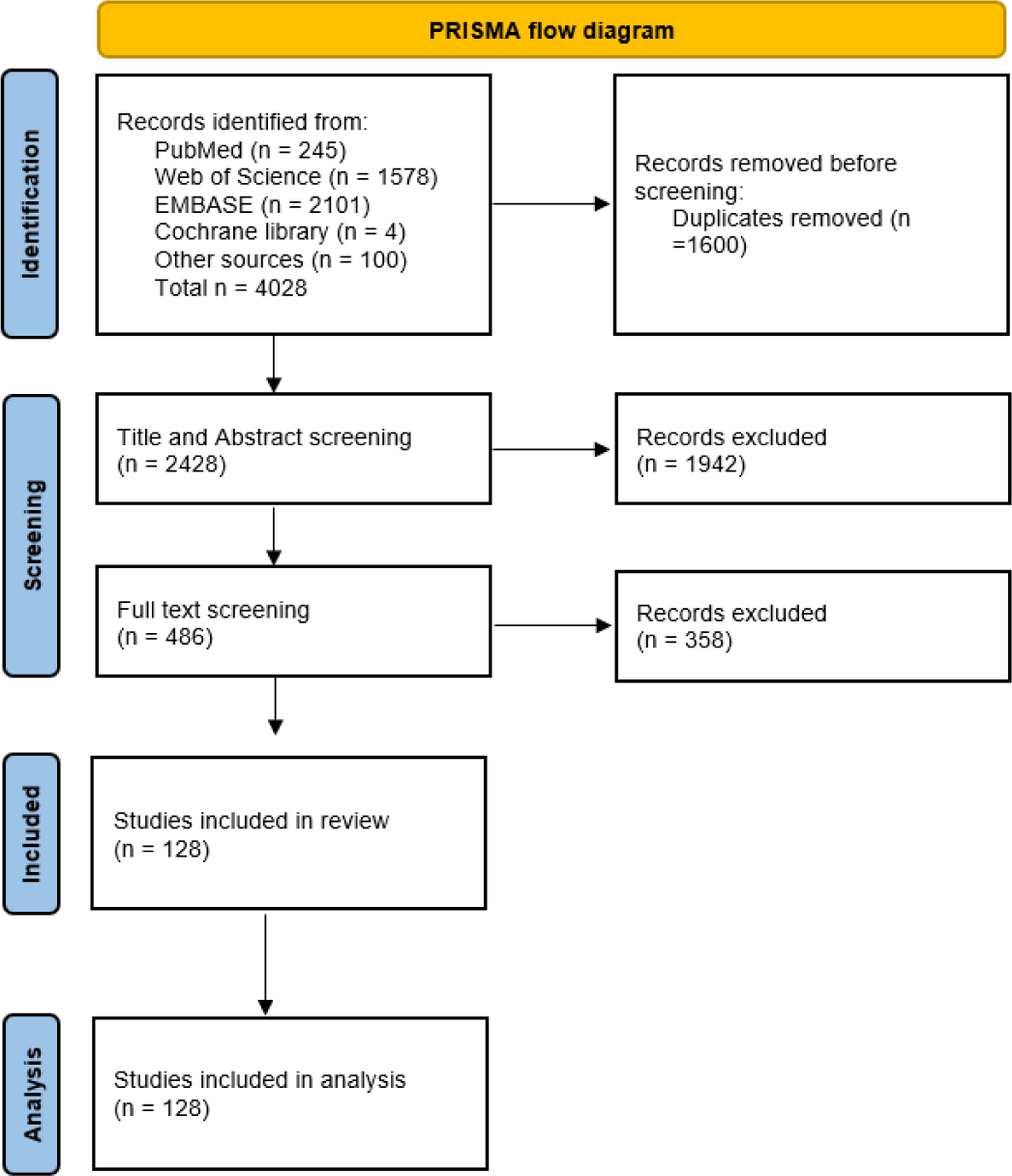
PRISMA flow diagram showing the studies included in the systematic review.

In total, the studies included 3057 patients of which 2441 (80%) were categorized as freezers either based on a clinical assessment by performing physical examinations or through self-reporting. 11599 FOG episodes were reported in 116 out of the 128 included studies. The mean age of all PD participants was 66.6 years ranging from 25.3 to 76.8 years. Furthermore, the average disease severity was determined based on Hoehn and Yahr, which resulted in disease stage 2.5, indicating mild bilateral disease [70]. Overall, more male participants (64%) were included in the studies than female participants (36%).

#### Patients with FOG

95 studies either reported the number or the percentage of their participants who experienced FOG throughout the respective gait trials. The patient cohort sample size, as well as the percentage of participants who experienced FOG, varied greatly between studies from 4 to 305 subjects and 0 to 100%, respectively (Figure 2). Looking at all participants with PD, 12 studies were not able to elicit any FOG in their test population, while 9 studies triggered FOG in all their participants. A total of 316 participants (average 26 participants per study) belongs to the group of studies with no FOG while the number of participants in studies with 100% FOG sums up to 121 (average 13). Out of the previously mentioned 80% of participants that were categorized as freezers, 54% experienced freezing in the course of the gait experiment.

**Figure 2.**
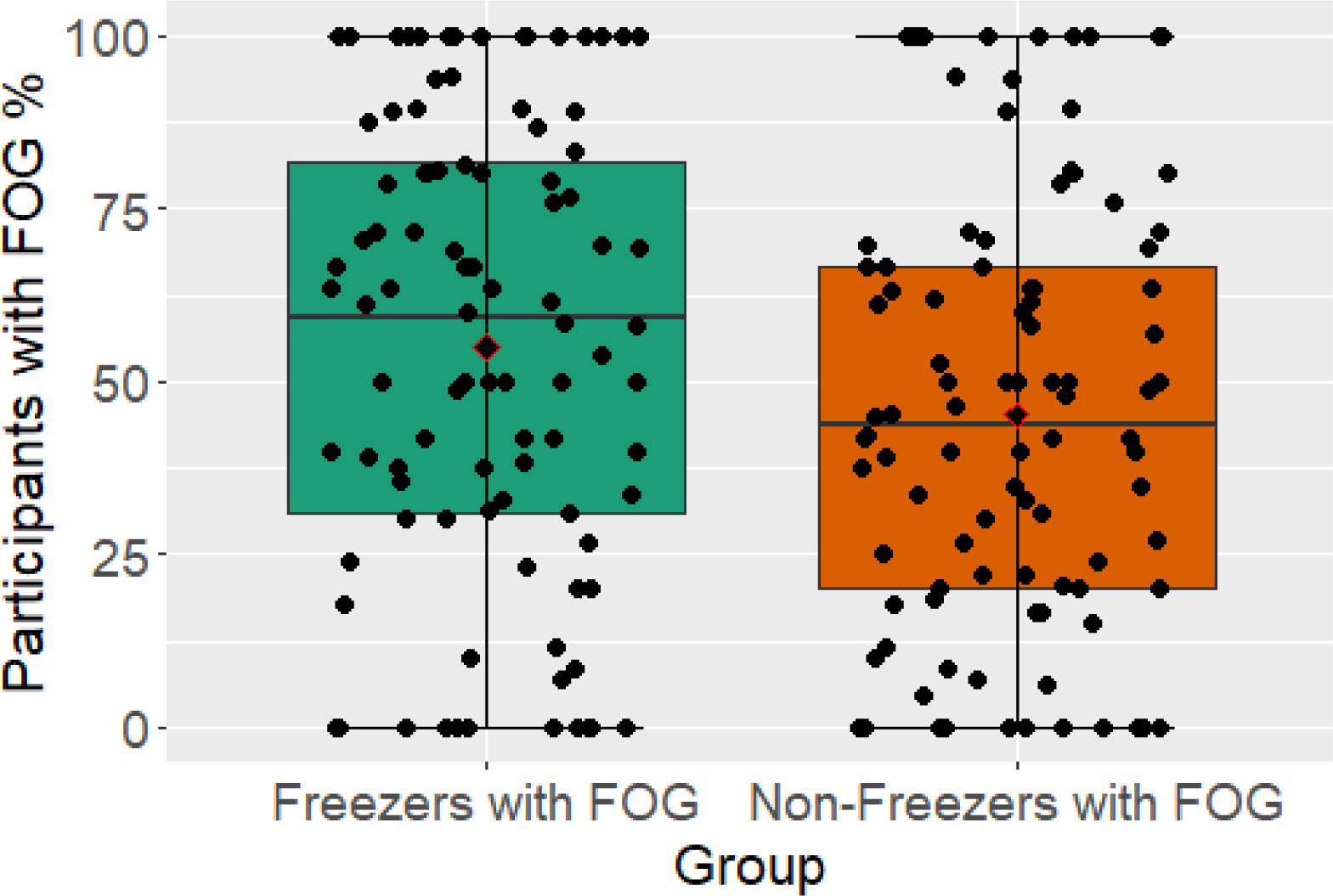
Boxplots showing the percentage of participants with FOG during gait trials for PwPD that are not known as freezers and the known freezers subgroup. Illustrated is the median with 25% and 75% confidence intervals.

#### Tasks

In total, 40 different gait tasks were reported and grouped into 21 categories based on higher-level tasks. Of the 128 included articles, only 22 studies exposed their participants to a single walking task, while the remainder requested their participants to perform at least two tasks. Turning of 180° (n = 38 studies) was the most used task, followed by 360° turning (n = 32 studies) (Figure 3). A few tasks were only conducted in a singular study, namely: long steps, augmented reality, turning of 120°, passing a wide door, and a *Six Minute Walking Test*. For the Turning and Barrier Course (TBC) subjects were “instructed to stand up, walk around the dividers twice in an ellipse, and then walk in a ‘figure eight’, around and through the opening between the dividers, twice, before sitting down again” [71]. Execution of the Ziegler course included a “Stand Up and Go test crossing through a doorway and then, turning back” [72]. Details on the study designs and execution of the other gait tasks can be found in the supplementary material (Supplementary Table 1).

**Figure 3.**
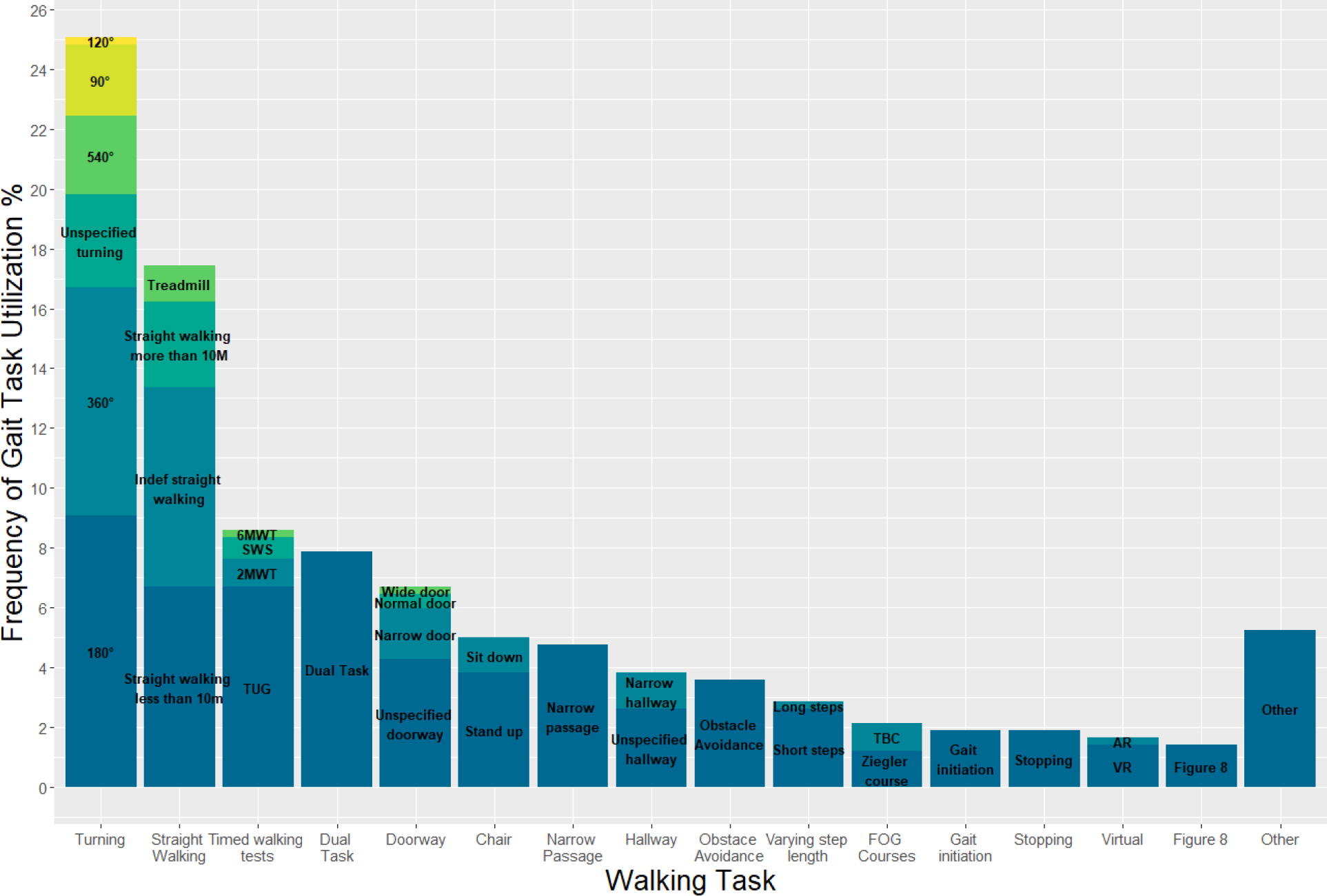
Stacked barplot displaying the different gait tasks used.

#### Trigger

26 of the included studies stated the gait task triggering FOG. This includes studies that examine participants in both ON- and OFF-medication condition. In total 24 different triggers were registered. Of these, the most effective trigger was 360° turning, which was responsible for 15.3% of FOG episodes recorded in the aforementioned studies (Figure 4). In addition, unspecified turning (10.1%), dual tasking (9.9%), stepping in place (9.5%), and passing a doorway (7.6%) were all activities with relatively high rates of eliciting FOG.

**Figure 4.**
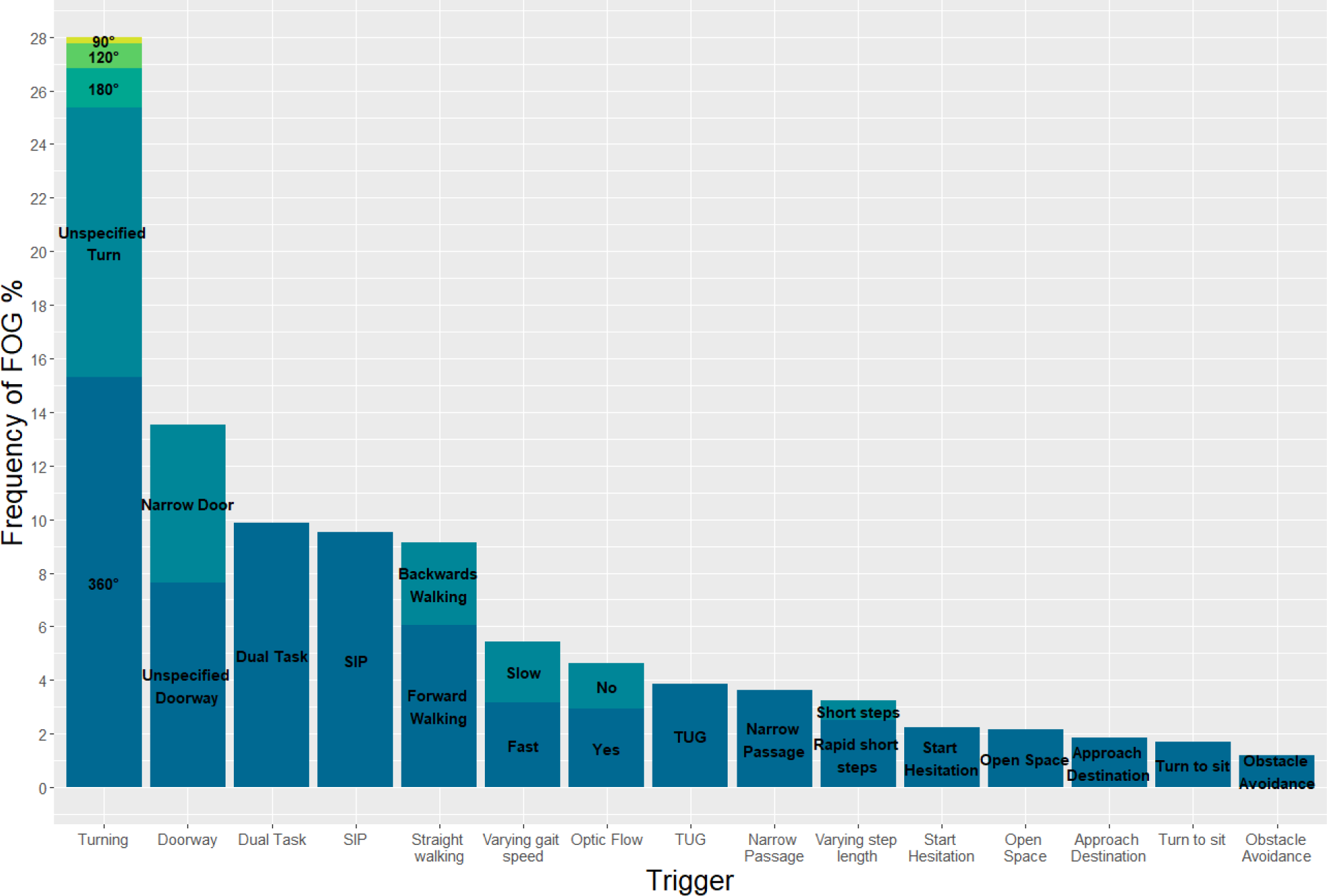
Stacked barplot demonstrating the triggers eliciting FOG in known freezers during gait trials.

## Discussion

Freezing of Gait is a debilitating symptom experienced by many people with Parkinson’s disease. Due to difficulties in eliciting FOG in a controlled laboratory or clinical setting, there has been a relative lack of research focused specifically on this phenomenon. Here, the challenge was to identify gait tasks that are most effective at triggering FOG. Therefore, a systematic review of the literature was conducted to provide valuable insights into the tasks that are most likely to elicit FOG, with the aim to support researchers in studying the underlying mechanisms of the symptom and develop effective interventions to improve the quality of life of PwPD. This systematic review provides evidence that turning is not only the most frequently studied gait task, but also the most effective gait task to induce FOG in PwPD (28%), followed by passing a doorway and dual tasking.

There are multiple theories surrounding the potent induction of FOG by turning. One hypothesis suggested by previous studies is that FOG is caused by a delay in maximum head-pelvis separation, leading to inadequate preparation for directional changes [32, 65]. Additionally, the asymmetrical stepping pattern induced by turning or the reduced ability of PwPD to adapt to a new gait pattern may also contribute to this phenomenon [73].

Cowie et al. (2012) reported a study that observed a slowing of walking in PwPD when approaching doorways, which they attributed to impaired visuomotor processing [55]. However, an alternative explanation could be that attention is diverted from walking when approaching a doorway, leading to reliance on automatic movement, which can be disrupted in some PwPD [20, 74]. Previous studies have demonstrated that dual tasking can lead to increased gait arrhythmicity and unsteadiness, as well as reduced step length and walking speed in PwPD [67, 75, 76]. This suggests that dual tasking may contribute to the occurrence of FOG, as patients divert their attention away from the gait task towards the secondary task, thereby increasing cognitive demand. Although several other gait tasks elicited FOG to a lesser extent, the diversity of triggers suggests that various brain areas and mechanisms are involved in the occurrence of FOG. Human movement involves a complex interplay of several brain areas [24, 77, 78], and any damage along the neural chain could therefore influence movement in different ways (Table 1) [24].

**Table 1.** Gait and possible neural mechanisms of different gait tasks.

| Task | Possible movement deficit | Possible neural deficit |
| --- | --- | --- |
| Turning | Re-orientation to a new direction [82]<br>Smaller step length on inner side of turning cycle than outside [31] | The basal ganglia provides phasic cues to the supplementary motor area (SMA), which regulates the bimanual coordination of movements [83]. The striatum is activated contralateral to the intended direction [84]. |
| Dual Task | Execution of a primary task (main focus of attention) while performing a secondary task simultaneously [85] | Cognitive prioritization of the executed tasks, the more attention-demanding task is controlled by frontal cortical areas [86]. The processing of the more automatic task (decreased dependency of attention) is shifted to the basal ganglia-circuit with input to the brainstem [20, 85]. |
| Obstacles<br>Doorways, narrow passages | Adjustment of ongoing locomotor patterns in order to adapt to changes in the environment [87] | Visual information in the visual cortex are translated to visuospatial information in the posterior parietal cortex (PPC) and motor output is generated by projections of the PPC to frontal motor regions. The visuospatial input and the motor output are compared online. The integration of proprioceptive and motor information might involve the basal ganglia [88]. |

In this systematic review, it was found that three gait tasks, namely turning, dual tasking, and straight walking, were performed in at least half of the studies analysed. Other gait tasks were performed much less frequently. The reasons for this preference for certain gait tasks can only be speculated on. It is possible that the choice of tasks was influenced by previous studies reporting successful triggering with a specific task or the ease of preparation and infrastructure required for the task. For instance, tests such as the TUG can be performed with minimal preparation, and adding a dual task to a pre-existing gait task can also be relatively simple.

A wide variation was observed in the percentage of participants experiencing FOG during gait assessment, ranging from 0% to 100% for both known freezers and PwPD (Figure 4). This variability is likely due to differences in study protocols, number of walking trials, medication status, and disease severity. However, the paroxysmal and unpredictable nature of FOG, influenced by environmental, emotional, and cognitive factors, is also a contributing factor [4, 26]. In the studies with no FOG episodes the most commonly examined task was straight walking. Nonetheless, all of the studies encompassed a diverse range of additional tasks, rendering it unfeasible to reach any definitive conclusions. In comparison to a previous study of almost a thousand participants, which reported a FOG prevalence of around one-third, the current study, involving a larger population of over 95 studies with exposure to a variety of tasks, showed a higher prevalence of around 50% for both PwPD and the subgroup of only Freezers [79]. This could enable a more enhanced exploration of the underlying mechanisms by the use of more effective triggers.

The male predominance in our sample is consistent with the fact that males are twice as likely to be affected by the disease than women [80]. The broad range of disease severity from I to almost IV further complicates comparison of study outcomes. Despite these limitations, this study provides a valuable contribution to the literature on FOG in PwPD and highlights the need for further investigation and standardization of FOG assessment protocols. With the study’s broad inclusion criteria, this is the first survey to comprehensively examine activities that elicit FOG by reviewing a large number of studies. However, the diverse nature of the studies included with varying execution of the conducted gait tasks posed a challenge in comparing outcomes. Additionally, a limitation of the study is the absence of a bias assessment to evaluate the quality of the included studies. Nonetheless, the study’s strengths, such as its comprehensive review and broad inclusion criteria, outweigh its limitations and contribute significantly to the current understanding of FOG in PwPD.

A noteworthy discovery from this study was the relatively low number of studies that focused on the triggers of FOG. The reason for this may be attributed to the challenge of accurately determining the cause of FOG episodes, which can be influenced by various emotional, environmental, and other factors. Interestingly, the frequency of FOG episodes did not significantly differ between the Freezers group and the entire PD patient population. This could be due to the unpredictable nature of FOG [10], or even the feeling of being observed and therefore increased attention while undertaking functional assessments in clinical and laboratory settings.[40].

In order to systematically study the underlying mechanisms of FOG, it is important to know which gait tasks act as the most efficient triggers for FOG. Current research is moving increasingly towards real-life assessment using IMUs [81]. However, this approach only informs about behavioural changes and does not provide information about the specific triggers that cause FOG. Therefore, more studies that focus specifically on the triggers of FOG are needed. This review provides evidence that turning is currently the most efficient trigger of FOG, but many different gait tasks are also able to elicit FOG to some extent. Based on these findings, turning might be the most effective task for FOG assessment in clinical examinations, but future studies should continue to cover a broad range of potential triggers in their protocol, as this can facilitate inference from phenomenological observation to underlying mechanisms. However, using different gait tasks separately according to a standardized protocol could simplify the process of precisely identifying and distinguishing the exact triggers of a FOG episode. Importantly, researchers should systematically annotate the cause of the FOG episode. This is particularly noteworthy as, until now, only a minority of studies explicitly report the actual triggers of FOG.

As the population continues to age, the prevalence of neurodegenerative diseases like PD is expected to rise. Thus, it is crucial to fully comprehend the symptoms, diagnosis, and treatment options associated with this condition. However, despite significant advancements in this area, research on the occurrence, severity, epidemiology, and underlying causes of FOG remains scarce. Therefore, this systematic review’s findings on triggers for FOG in PwPD will be valuable in guiding future research and clinical applications, aiding in the selection of assessments for FOG. Emerging technologies such as augmented or virtual reality hold promise in this field, as they can be utilized to evaluate, diagnose, and cue FOG effectively. These technologies can incorporate individual triggers as building blocks, allowing for the creation of personalized walking courses tailored to an individual’s preferred difficulty level.

## Conclusion

This review offers a significant contribution to the understanding of FOG triggers in PwPD by providing a comprehensive overview. The results indicate that turning is the most effective trigger for FOG in PwPD, followed by walking through a doorway and dual tasking. These findings have potential applications for researchers designing studies on FOG, clinicians evaluating patients with PD, and the development of interventions to manage or prevent FOG. Implementing the results to design coherent recommendations in research as well as clinical evaluation can lead to a better understanding of FOG and therefore an improvement of current treatment.

## Data Availability

The data supporting the findings of this study are available within the article and/or its supplementary material.

## Ethical Compliance Statement

The authors confirm that the approval of an institutional review board was not required for this work. Informed patient consent was not necessary for this work.

## Acknowledgement

We thank for Pino Wüest for his assistance in data collection.

## Supplementary material

**Supplementary methods 1** Complete search string

**Supplementary table 1** Extracted information of studies included

