## Supplementary_methods_1 for "Triggers for Freezing of Gait in Individuals with Parkinson’s Disease: A Systematic Review"

**Complete search string**

(parkin* OR PD OR "Parkinson's disease" OR "Parkinsonian disorders" OR "Morbus parkinson" OR “shaking palsy”)

AND

("Freezing of gait" OR FOG)

AND

(gait OR walking OR walk OR locomotion OR locomotor OR shuffling OR festination)

NOT

(child* OR animal* OR gene OR cell OR "Nucleic acid" OR "Genome components" OR acid OR oxidation OR nanoparticles OR DNA OR RNA OR children OR rat OR mice OR monkey OR coli OR cats OR fishes OR robot* OR amput* OR arm OR hand OR cryo* OR bacteria OR virus OR meat)
